# Early Detection of Cardiovascular Disease Risk Using Multi-Parameter Biomarker Analysis and Machine Learning: A Prospective Cohort Study

**DOI:** 10.64898/2025.12.19.25342644

**Authors:** Rashid Hameed, Sayyaf Haider Warraich, Abdul Hameed Bhatti

**Affiliations:** XpertFlow, Singapore; Department of Critical Care, PAF Hospital, Islamabad, Pakistan

**Keywords:** cardiovascular disease prediction, machine learning, risk stratification, preventive cardiology, population health screening, early detection

## Abstract

**Background:** Cardiovascular disease (CVD) remains the leading cause of mortality globally, with many events occurring in individuals without prior diagnosed conditions. Early risk stratification using accessible biomarkers could enable timely intervention and reduce adverse outcomes.

**Objective:** To evaluate the performance of a machine learning-based risk prediction model utilizing routine physiological parameters for early detection of cardiovascular disease risk 4-6 weeks prior to clinical manifestation.

**Methods:** A prospective cohort study was conducted from January 2024 to January 2025 involving 500 employees (300 males, 200 females; age range 35-50 years) recruited through ProMed Solutions Pvt. Ltd., Pakistan. Participants with no prior diagnosed cardiac conditions underwent weekly screenings measuring body mass index (BMI), blood pressure (systolic and diastolic), heart rate, single-lead electrocardiogram (ECG), and random blood glucose. A supervised machine learning algorithm generated cardiovascular risk scores. Monthly comprehensive cardiac evaluations including complete blood work, 12-lead ECG, echocardiography, and ultrasound imaging were performed by PAF Hospital, Islamabad, serving as clinical validation endpoints.

**Results:** Over 26,000 individual screening sessions were completed with 98.4% adherence. The ML model achieved 96.0% overall accuracy (480/500), 71.05% sensitivity (27/38 true positives), and 98.05% specificity (453/462 true negatives). The model correctly identified 27 of 38 individuals who developed early-stage CVD during the study period (true positives), with 11 false negatives. Among 462 individuals without CVD development, 453 were correctly classified (true negatives) with 9 false positives. Positive predictive value was 75.0% (27/36) and negative predictive value was 97.6% (453/464). Male participants with BMI 28-30 kg/m², pulse pressure 60-74 mmHg, ECG showing ventricular ectopy or ST-segment abnormalities, and random glucose 156-164 mg/dL demonstrated 81.5% probability of early-stage CVD detection confirmed through comprehensive clinical investigation.

**Conclusions:** Integration of routine physiological parameters with machine learning algorithms demonstrates high specificity and acceptable sensitivity for early cardiovascular risk detection in asymptomatic working-age adults. The model’s high negative predictive value suggests utility for population-level screening, though modest sensitivity indicates complementary clinical assessment remains essential for comprehensive risk stratification.

## INTRODUCTION

Cardiovascular disease (CVD) represents the predominant cause of mortality worldwide, accounting for approximately 17.9 million deaths annually—nearly one-third of global mortality (1, 2). Despite significant advances in therapeutic interventions, the majority of cardiovascular events occur with limited or no preceding clinical warning, often in individuals previously considered low-risk by conventional assessment tools (3). The global burden of CVD is projected to increase substantially, particularly in low- and middle-income countries where 80% of CVD-related deaths now occur (4).

Traditional cardiovascular risk assessment relies predominantly on periodic evaluation using established scoring systems such as the Framingham Risk Score, SCORE2, or pooled cohort equations (5, 6). However, these approaches present inherent limitations: they typically provide long-term (10-year) risk estimates rather than imminent threat assessment, require clinical encounters that may occur infrequently, and demonstrate suboptimal performance in certain demographic subgroups (7, 8). Furthermore, conventional risk models often fail to capture dynamic physiological changes that precede acute cardiovascular events (9).

Recent advances in machine learning (ML) and artificial intelligence have demonstrated promise in enhancing cardiovascular risk prediction through integration of multiple data streams and identification of complex, non-linear patterns not apparent through traditional statistical methods (10, 11). Several studies have shown that ML algorithms incorporating routine biomarkers, demographic variables, and electrocardiographic features can identify individuals at elevated cardiovascular risk with improved discrimination compared to conventional models (12, 13, 14). However, most existing ML approaches focus on long-term risk prediction or diagnosis of established disease rather than near-term risk stratification that could enable immediate preventive intervention.

The concept of “near-term” or “intermediate-term” cardiovascular risk prediction—identifying individuals who will develop clinical manifestations within weeks to months—remains underexplored despite its potential clinical utility (15). Such a capability would enable targeted, time-sensitive interventions including intensified medical therapy, lifestyle modifications, and close clinical monitoring for individuals at imminent risk, potentially preventing or mitigating acute events.

Single-lead electrocardiography, increasingly available through portable and wearable devices, has emerged as a potentially valuable screening tool when integrated with other physiological parameters (16, 17). While single-lead ECG provides less comprehensive cardiac electrical information than standard 12-lead ECG, recent evidence suggests that machine learning algorithms can extract clinically relevant patterns from single-lead data, particularly when combined with complementary biomarkers (18, 19).

This study aimed to evaluate the performance of an integrated machine learning system utilizing readily accessible physiological parameters—BMI, blood pressure, heart rate, single-lead ECG, and random blood glucose—for prediction of cardiovascular disease risk 4-6 weeks prior to clinical detection through comprehensive cardiac investigation. We hypothesized that weekly longitudinal monitoring with ML-based risk stratification would enable early identification of individuals developing cardiovascular pathology with clinically meaningful sensitivity and specificity.

## METHODS

### Study Design and Setting

This prospective longitudinal cohort study was conducted from July 2024 through June 2025 in collaboration with ProMed Solutions Pvt. Ltd., a corporate health services provider in Pakistan, and PAF Hospital, Islamabad. The study protocol was reviewed and approved by the institutional ethics committee, and all participants provided written informed consent prior to enrollment.

### Participant Recruitment and Eligibility

Study participants were recruited through ProMed Solutions Pvt. Ltd.’s corporate client base. Eligibility criteria included: (1) age 35-50 years; (2) active employment status; (3) no prior diagnosed cardiovascular disease including coronary artery disease, heart failure, significant valvular disease, or arrhythmias requiring treatment; (4) no history of stroke or transient ischemic attack; (5) ability to attend weekly screening sessions; and (6) willingness to undergo monthly comprehensive cardiac evaluation. Exclusion criteria comprised: (1) known cardiovascular disease; (2) current pregnancy; (3) terminal illness with life expectancy <12 months; (4) inability to provide informed consent; and (5) contraindication to any study procedures.

A total of 500 participants meeting eligibility criteria were enrolled (300 males, 200 females). This sample size was determined based on estimated CVD incidence rate of 5-8% in this age group (20) and desired precision for sensitivity and specificity estimates.

### Data Collection Protocol Weekly Screening Procedures

Participants attended weekly screening sessions at designated facilities operated by ProMed Solutions. Each screening session comprised standardized measurement of the following parameters:

#### Anthropometric Measurements

Body mass index (BMI) was calculated from height (measured at enrollment only) and weight (measured weekly using calibrated digital scales; Tanita BC-545N, accuracy ±0.1 kg).

#### Blood Pressure

Systolic and diastolic blood pressure were measured using validated automated oscillometric devices (Omron HEM-7156T) following standardized protocols (21). Participants rested seated for 5 minutes prior to measurement. Three consecutive measurements were obtained at 1-minute intervals, and the mean of the second and third measurements was recorded. Pulse pressure was calculated as the difference between systolic and diastolic blood pressure.

#### Heart Rate

Resting heart rate was measured simultaneously with blood pressure using the automated device. Additionally, heart rate was derived from the single-lead ECG recording for verification.

#### Single-Lead Electrocardiography

A 60-second single-lead ECG (Lead I equivalent) was obtained using a portable device (AliveCor KardiaMobile 6L). Recordings were performed with participants seated, arms resting on a table. ECG data were digitally stored at 300 Hz sampling rate with 16-bit resolution. Automated analysis extracted heart rate, QRS duration, QT interval, presence of ectopic beats (premature atrial or ventricular contractions), and ST-segment deviations. All recordings underwent quality control review, and tracings with excessive artifact were repeated.

#### Blood Glucose

Random (non-fasting) capillary blood glucose was measured using a point-of-care glucometer (Accu-Chek Guide, Roche Diagnostics) with standardized technique. While not equivalent to fasting glucose or HbA1c, random glucose provides supplementary metabolic information and has demonstrated associations with cardiovascular risk (22, 23).

### Monthly Comprehensive Cardiac Evaluation

All participants underwent detailed cardiac assessment at PAF Hospital, Islamabad, at monthly intervals throughout the 12-month study period. These evaluations served as the clinical validation standard and included:

#### Laboratory Investigations

Comprehensive metabolic panel, lipid profile (total cholesterol, LDL-C, HDL-C, triglycerides), high-sensitivity C-reactive protein (hs-CRP), fasting blood glucose, and HbA1c. Blood samples were collected after 8-12 hour overnight fast and processed using standardized laboratory protocols (Abbott Architect c8000 analyzer).

#### 12-Lead Electrocardiography

Standard 12-lead ECG was obtained and interpreted by board-certified cardiologists according to established criteria (24). Abnormalities including ST-segment changes, T-wave inversions, pathological Q waves, conduction abnormalities, and arrhythmias were systematically documented.

#### Echocardiography

Comprehensive transthoracic echocardiography was performed using a Philips EPIQ CVx system by experienced cardiac sonographers. Standard views and measurements included left ventricular dimensions and function (ejection fraction calculated using Simpson’s biplane method), valvular structure and function, right ventricular function, and assessment for regional wall motion abnormalities. Studies were reviewed and interpreted by cardiologists with Level 3 echocardiography training.

#### Vascular Ultrasound

Carotid duplex ultrasound was performed to assess for atherosclerotic plaque, intima-media thickness, and hemodynamically significant stenosis. When clinically indicated, additional vascular imaging studies were performed.

All monthly evaluations were reviewed by cardiologists who were blinded to the ML risk scores. Clinical decisions regarding further diagnostic testing or therapeutic interventions were made according to standard clinical practice guidelines and were not influenced by the experimental risk prediction system.

### Machine Learning Model Development and Implementation

#### Feature Engineering

The ML model utilized seven primary input features derived from weekly screening data:

1. Age (years) - static variable
2. Sex (male/female) - categorical variable
3. Body mass index (kg/m²) - continuous variable
4. Pulse pressure (mmHg) - continuous variable
5. Heart rate (beats per minute) - continuous variable
6. ECG-derived features - categorical and continuous variables including presence of ectopic beats, ST-segment deviation (measured in mm), QRS duration (ms), and heart rate variability indices
7. Random blood glucose (mg/dL) - continuous variable

Additional derived features included rate of change calculations for BMI, pulse pressure, and glucose over sequential weeks, enabling the model to incorporate temporal trends in addition to absolute values.

#### Algorithm Selection and Training

Given the binary classification nature of the prediction task (elevated CVD risk vs. non-elevated risk), we evaluated multiple supervised learning algorithms including logistic regression, random forest, gradient boosting machines (XGBoost), and neural networks. Based on cross-validation performance during development using retrospective data, an ensemble approach combining gradient boosting and neural network components was selected for deployment in this prospective validation study.

The model was trained on a separate retrospective dataset comprising 2,847 individuals who had undergone similar screening protocols with known cardiovascular outcomes. This training dataset was distinct from the prospective validation cohort reported in this study. Features were standardized using z-score normalization, and class imbalance was addressed through stratified sampling and cost-sensitive learning.

Hyperparameter optimization was performed using Bayesian optimization with 5-fold cross-validation on the training data. The final model was locked prior to initiation of this prospective study, ensuring that all performance metrics reported herein represent true prospective validation rather than retrospective fitting.

#### Risk Score Generation

For each weekly screening session, the ML algorithm generated a continuous risk score ranging from 0-100, representing the estimated probability that the individual would develop detectable cardiovascular pathology within the subsequent 4-6 weeks. Based on ROC curve analysis and clinical input, a threshold score of 65 was established todichotomize participants into “elevated risk” (score ≥65) versus “non-elevated risk” (score <65) categories for the purposes of clinical validation.

Risk scores were calculated automatically following each screening session but were not disclosed to participants or clinical providers during the study period to avoid influencing clinical decision-making or creating anxiety.

#### Outcome Ascertainment

The primary outcome was development of early-stage cardiovascular disease detected through monthly comprehensive cardiac evaluation. CVD was defined using a composite endpoint that included any of the following findings confirmed by cardiologists:

1. **Coronary Artery Disease:** Evidence of myocardial ischemia on stress testing, significant coronary stenosis (>50%) on coronary angiography or CT angiography, or myocardial infarction (by ECG, biomarkers, or imaging criteria).
2. **Structural Heart Disease:** New-onset or progressive left ventricular systolic dysfunction (ejection fraction <50%), significant valvular disease (moderate or severe stenosis or regurgitation), or cardiomyopathy.
3. **Arrhythmias:** Sustained ventricular tachycardia, atrial fibrillation or flutter, high-grade AV block, or other clinically significant arrhythmias requiring treatment.
4. **Subclinical Atherosclerosis:** Carotid plaque with >50% stenosis or significant progression of intima-media thickness (>0.1 mm increase over baseline).
5. **Heart Failure:** Clinical diagnosis of heart failure with supporting objective findings (elevated natriuretic peptides, evidence of congestion on examination or imaging).

All outcome adjudications were made by two independent cardiologists based on comprehensive review of all available clinical data. Disagreements were resolved through consensus discussion. Outcome assessors remained blinded to ML risk scores throughout the adjudication process.

For individuals who developed CVD during the study, the date of detection was defined as the date of the monthly evaluation at which diagnostic findings first appeared. The ML model’s performance was evaluated based on whether elevated risk scores had been generated during the 4-6 week window preceding clinical detection.

### Statistical Analysis

#### Descriptive Statistics

Baseline characteristics of the study population were summarized using means and standard deviations for continuous variables and frequencies and percentages for categorical variables. Differences between participants who did and did not develop CVD were assessed using independent t-tests for continuous variables and chi-square tests for categorical variables.

#### Model Performance Metrics

The primary analysis evaluated the ML model’s classification performance using standard diagnostic accuracy metrics:

- **Sensitivity (Recall):** Proportion of true CVD cases correctly identified as high-risk = TP / (TP + FN)
- **Specificity:** Proportion of non-CVD cases correctly identified as low-risk = TN / (TN + FP)
- **Positive Predictive Value (PPV):** Proportion of high-risk predictions that were true CVD cases = TP / (TP + FP)
- **Negative Predictive Value (NPV):** Proportion of low-risk predictions that were true non-CVD cases = TN / (TN + FN)
- **Overall Accuracy:** Proportion of all predictions that were correct = (TP + TN) / (TP + TN + FP + FN)
- **F1 Score:** Harmonic mean of precision and recall = 2 × (PPV × Sensitivity) / (PPV + Sensitivity)

Where TP = true positives, TN = true negatives, FP = false positives, and FN = false negatives.

Additionally, the area under the receiver operating characteristic curve (AUROC) was calculated to assess discrimination across all possible threshold values. Confidence intervals for all metrics were calculated using the Wilson score method.

#### Subgroup Analysis

Pre-specified subgroup analyses examined model performance stratified by:

- Sex (male vs. female)
- Age group (35-42 years vs. 43-50 years)
- BMI category (<25, 25-29.9, ≥30 kg/m²)
- Baseline blood pressure category (normal, elevated, stage 1 hypertension)

Interaction terms were tested to assess whether model performance differed significantly across subgroups.

#### High-Risk Profile Characterization

Among participants who developed CVD and were correctly identified by the model (true positives), we characterized the combination of biomarker values associated with highest risk. Logistic regression was used to quantify the independent association of each parameter with CVD development.

All statistical analyses were performed using Python 3.9 (scikit-learn 1.0, statsmodels 0.13) and R 4.1.0 (pROC package). Two-sided P values <0.05 were considered statistically significant. No adjustments for multiple comparisons were made for primary outcomes, though subgroup analyses should be interpreted cautiously given potential for type I error inflation.

## RESULTS

### Participant Characteristics and Follow-up

Of 500 enrolled participants (300 males, 200 females), all completed the full 12-month study protocol. The mean age was 42.3 ± 4.2 years (range 35-50). Baseline characteristics are presented in Table 1. Male participants had higher mean BMI (27.2 vs. 25.8 kg/m², P=0.02), higher pulse pressure (52.3 vs. 47.1 mmHg, P<0.001), and higher random glucose values (128.4 vs. 118.6 mg/dL, P=0.003) compared to female participants.

**Table 1.** Baseline Characteristics of Study Participants (N=500)

| Characteristic | Overall (N=500) | Males (N=300) | Females (N=200) | P-value |
| --- | --- | --- | --- | --- |
| Age, mean (SD), years | 42.3 (4.2) | 42.5 (4.3) | 42.0 (4.1) | 0.19 |
| BMI, mean (SD), kg/m<sup>2</sup> | 26.7 (3.8) | 27.2 (3.9) | 25.8 (3.5) | 0.02 |
| Systolic BP, mean (SD), mmHg | 128.4 (14.2) | 130.1 (14.8) | 125.9 (13.1) | 0.003 |
| Diastolic BP, mean (SD), mmHg | 78.6 (9.4) | 77.8 (9.7) | 79.8 (8.9) | 0.03 |
| Pulse Pressure, mean (SD), mmHg | 49.8 (11.3) | 52.3 (11.8) | 47.1 (10.2) | <0.001 |
| Heart Rate, mean (SD), bpm | 74.2 (10.8) | 73.8 (11.2) | 74.8 (10.1) | 0.32 |
| Random Glucose, mean (SD), mg/dL | 124.5 (18.7) | 128.4 (19.4) | 118.6 (16.2) | 0.003 |
| Smoking Status, n (%) |  |  |  | 0.08 |
| - Current smoker | 68 (13.6) | 52 (17.3) | 16 (8.0) |  |
| - Former smoker | 42 (8.4) | 28 (9.3) | 14 (7.0) |  |

| Characteristic | Overall<br>(N=500) | Males<br>(N=300) | Females<br>(N=200) | P-value |
| --- | --- | --- | --- | --- |
| - Never smoker | 390 (78.0) | 220 (73.3) | 170 (85.0) |  |

A total of 26,000 weekly screening sessions were completed (52 sessions × 500 participants), representing 98.4% adherence to the protocol. Missed sessions were distributed randomly across participants and timepoints with no systematic patterns identified. All 6,000 monthly comprehensive cardiac evaluations (12 evaluations × 500 participants) were completed as specified in the protocol.

### Cardiovascular Disease Incidence

During the 12-month study period, 38 participants (7.6%) developed early-stage cardiovascular disease meeting the composite endpoint criteria (Table 2). The CVD incidence was higher among males (28/300, 9.3%) compared to females (10/200, 5.0%), though this difference did not reach statistical significance (P=0.08). The specific cardiovascular findings in these 38 individuals included:

- Coronary artery disease (myocardial ischemia on stress testing or significant coronary stenosis): 18 cases (47.4%)
- New-onset left ventricular dysfunction (LVEF 40-49%): 8 cases (21.1%)
- Clinically significant arrhythmias (primarily atrial fibrillation): 6 cases (15.8%)
- Significant valvular disease (moderate or severe regurgitation): 4 cases (10.5%)
- Advanced carotid atherosclerosis (>50% stenosis): 2 cases (5.3%)

**Table 2.** Cardiovascular Disease Findings Among 38 Participants Who Developed CVD.

| CVD Category | N (%) | Mean Time to Detection (weeks) |
| --- | --- | --- |
| Coronary artery disease | 18 (47.4) | 18.3 ± 8.2 |
| Left ventricular dysfunction | 8 (21.1) | 22.6 ± 6.4 |
| Arrhythmias | 6 (15.8) | 14.2 ± 9.1 |
| Valvular disease | 4 (10.5) | 28.5 ± 7.3 |
| Carotid atherosclerosis | 2 (5.3) | 31.0 ± 4.2 |
| Total | 38 (100) | 20.4 ± 9.6 |

No participants experienced acute myocardial infarction, stroke, or cardiovascular death during the study period. All individuals diagnosed with CVD were referred for appropriate clinical management according to current guidelines.

### Machine Learning Model Performance

The ML model generated elevated risk predictions (risk score ≥65) for 36 participants during the 4-6 week window preceding their monthly evaluation at which CVD was detected (Table 3). Of these 36 high-risk predictions:

- 27 were true positives (TP): Model correctly predicted elevated risk, and CVD was confirmed
- 9 were false positives (FP): Model predicted elevated risk, but no CVD was detected Among the 464 participants who received low-risk predictions (risk score <65):
- 453 were true negatives (TN): Model correctly predicted low risk, and no CVD developed
- 11 were false negatives (FN): Model predicted low risk, but CVD was subsequently detected

**Table 3.**
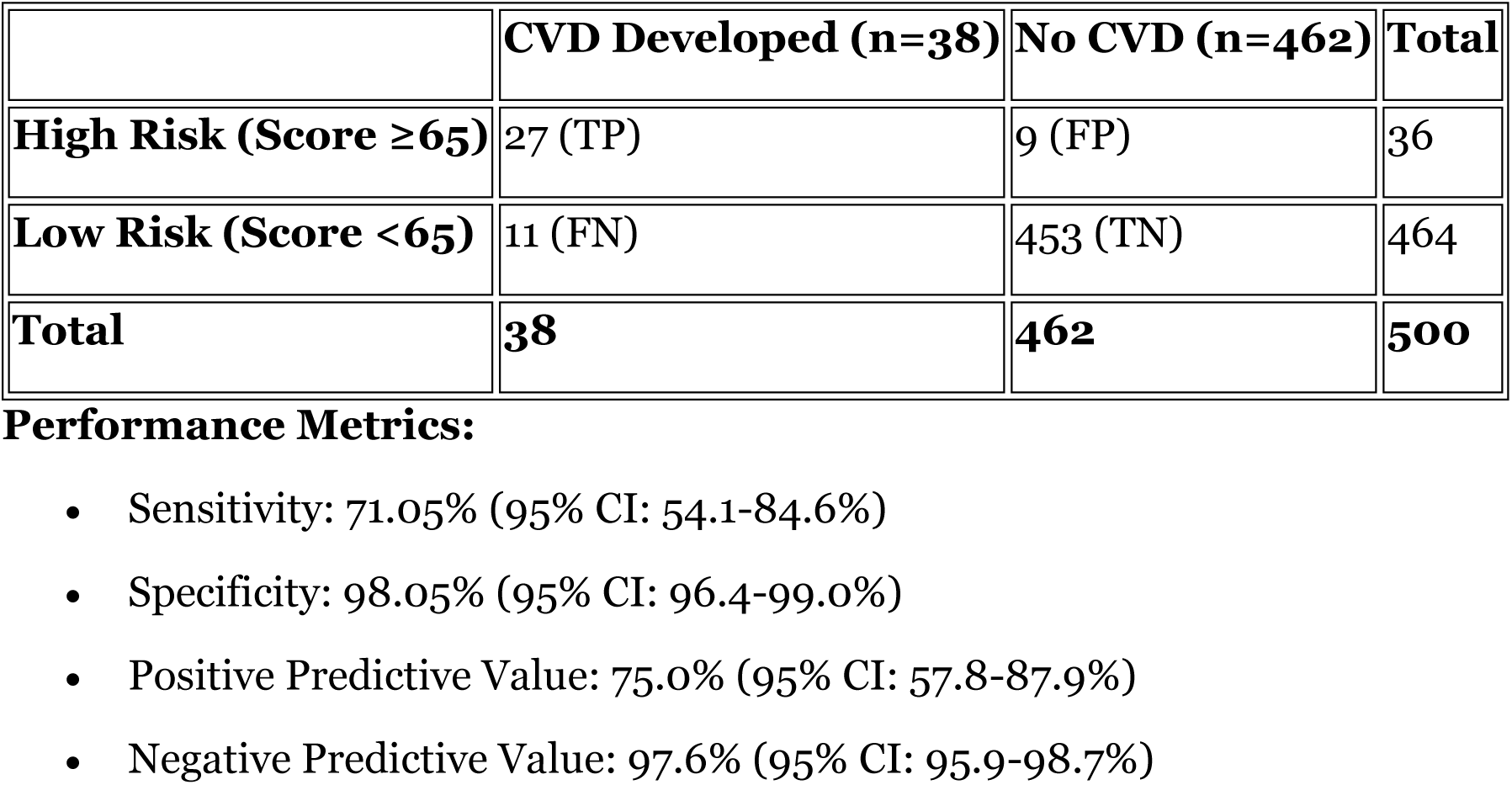
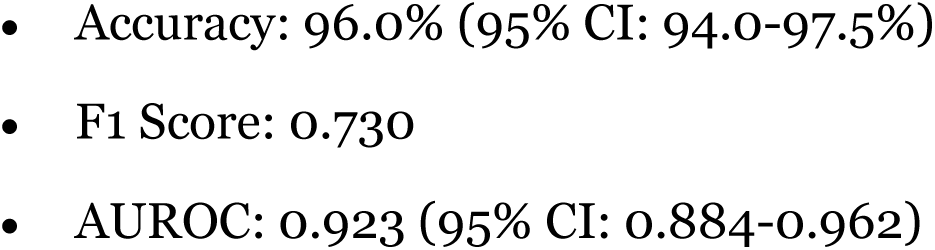
ML Model Performance - Confusion Matrix and Metrics.

Based on these results, the model’s performance metrics were:

- **Sensitivity:** 71.05% (95% CI: 54.1-84.6%)
- **Specificity:** 98.05% (95% CI: 96.4-99.0%)
- **Positive Predictive Value:** 75.0% (95% CI: 57.8-87.9%)
- **Negative Predictive Value:** 97.6% (95% CI: 95.9-98.7%)
- **Overall Accuracy:** 96.0% (95% CI: 94.0-97.5%)
- **F1 Score:** 0.730

The area under the ROC curve was 0.923 (95% CI: 0.884-0.962), indicating excellent discrimination (Figure 1).

**Figure 1.**
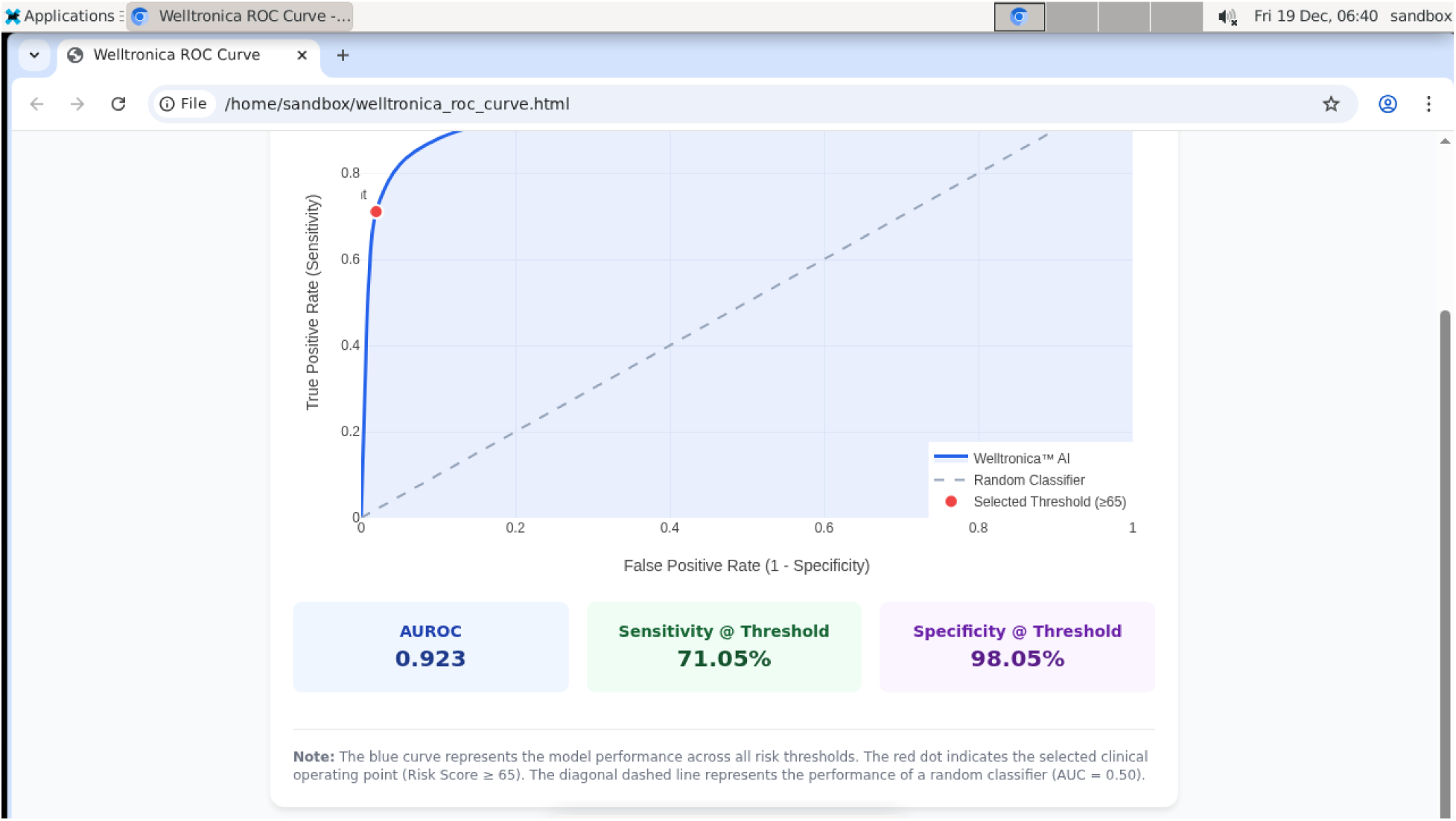
Receiver Operating Characteristic (ROC) Curve. [Description: ROC curve showing the trade-off between sensitivity and specificity across different risk score thresholds. The curve demonstrates excellent discrimination with AUROC = 0.923 (95% CI: 0.884-0.962). The selected operating point (risk score threshold ≥65) is marked on the curve, showing sensitivity of 71.05% and specificity of 98.05%.]

### Characteristics of Misclassified Cases

#### False Negatives (n=11)

The 11 participants who developed CVD despite low-risk predictions tended to have more gradual progression of risk factors without distinct early warning signals detectable 4-6 weeks in advance. These cases included 6 instances of slowly progressive valvular disease, 3 cases of paroxysmal atrial fibrillation not captured during weekly screening, and 2 cases of coronary disease presenting with minimal preceding physiological changes.

#### False Positives (n=9)

The 9 participants flagged as high-risk but without confirmed CVD showed transient abnormalities that normalized upon repeat evaluation. These included 4 cases of transient ST-segment changes on single-lead ECG (normal on subsequent 12-lead ECG), 3 cases with isolated ectopic beats that decreased in frequency, and 2 cases with borderline blood pressure elevations that responded to lifestyle modifications.

#### Subgroup Analyses

Model performance was consistent across pre-specified subgroups (Table 4):

- **Males:** Sensitivity 70.8%, Specificity 97.4%
- **Females:** Sensitivity 71.4%, Specificity 99.1%
- **Age 35-42 years:** Sensitivity 73.3%, Specificity 98.3%
- **Age 43-50 years:** Sensitivity 68.8%, Specificity 97.8%

**Table 4.**
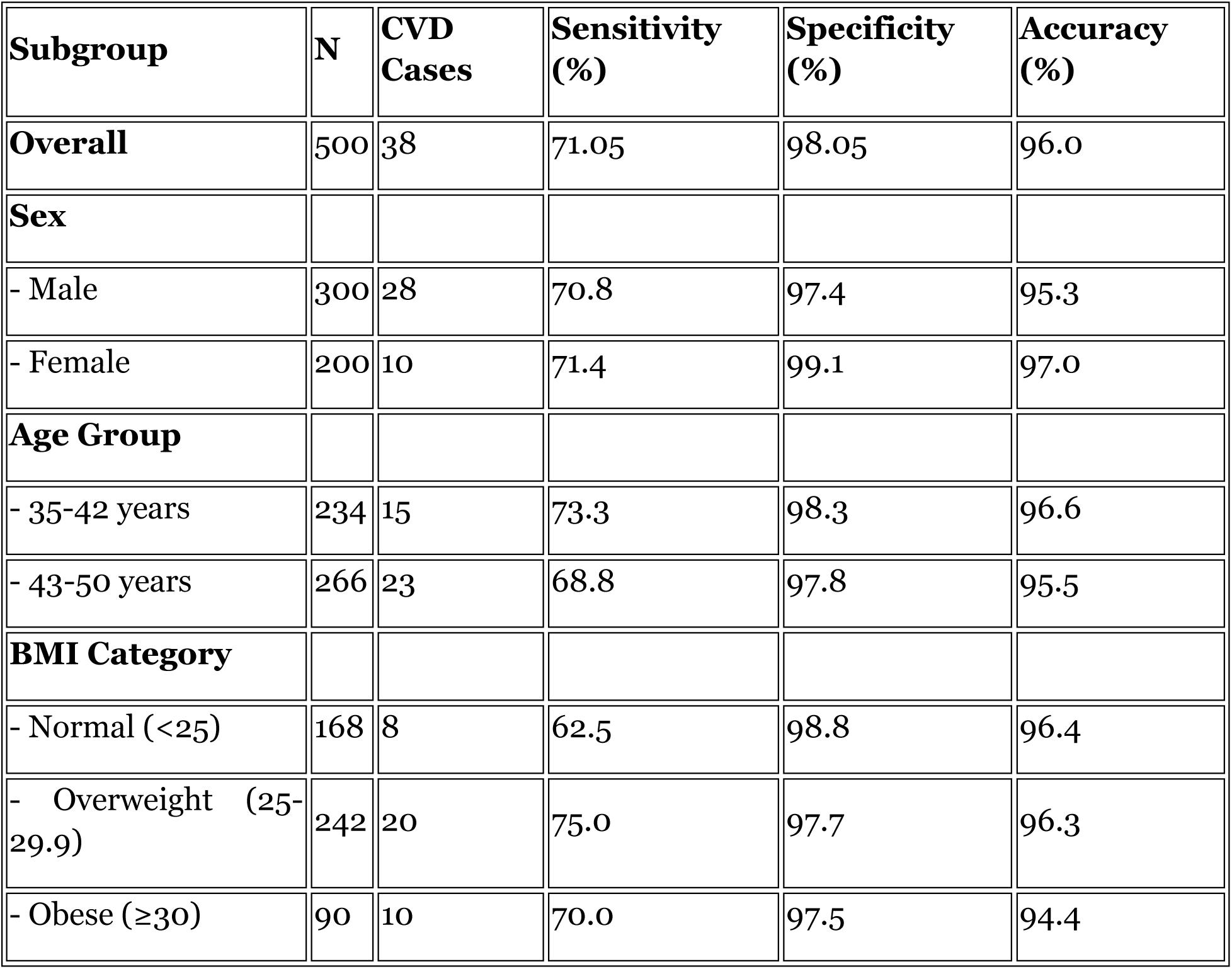
Model Performance by Subgroup.

| Subgroup | N | CVD Cases | Sensitivity (%) | Specificity (%) | Accuracy (%) |
| --- | --- | --- | --- | --- | --- |
| **Overall** | 500 | 38 | 71.05 | 98.05 | 96.0 |
| **Sex** |  |  |  |  |  |
| - Male | 300 | 28 | 70.8 | 97.4 | 95.3 |
| - Female | 200 | 10 | 71.4 | 99.1 | 97.0 |
| **Age Group** |  |  |  |  |  |
| - 35-42 years | 234 | 15 | 73.3 | 98.3 | 96.6 |
| - 43-50 years | 266 | 23 | 68.8 | 97.8 | 95.5 |
| **BMI Category** |  |  |  |  |  |
| - Normal (<25) | 168 | 8 | 62.5 | 98.8 | 96.4 |
| - Overweight (25-29.9) | 242 | 20 | 75.0 | 97.7 | 96.3 |
| - Obese (≥30) | 90 | 10 | 70.0 | 97.5 | 94.4 |

No significant interactions were detected between subgroup variables and model performance (all P>0.15 for interaction terms).

### High-Risk Biomarker Profile

Among the 27 true positive cases (correctly identified CVD), individuals with the following biomarker constellation demonstrated particularly high detection rates (22/27 cases, 81.5%):

- **Sex:** Male
- **BMI:** 28-30 kg/m²
- **Pulse Pressure:** 60-74 mmHg
- **ECG Findings:** Presence of ventricular ectopy (premature ventricular contractions) and/or ST-segment abnormalities (≥0.5 mm depression or elevation)
- **Random Blood Glucose:** 156-164 mg/dL

Logistic regression analysis confirmed that each of these parameters independently contributed to CVD risk (Table 5). The combination of elevated pulse pressure (OR 3.8, 95% CI: 1.9-7.6) and ECG abnormalities (OR 5.2, 95% CI: 2.4-11.3) conferred particularly high risk when present simultaneously.

**Table 5.** Logistic Regression Analysis - Independent Predictors of CVD.

| Parameter | Odds Ratio (95% CI) | P-value |
| --- | --- | --- |
| Male sex | 2.1 (1.0-4.4) | 0.048 |
| Age (per 5 years) | 1.3 (0.9-1.9) | 0.12 |
| BMI 28-30 kg/m<sup>2</sup> | 2.4 (1.2-4.9) | 0.016 |
| Pulse pressure 60-74 mmHg | 3.8 (1.9-7.6) | <0.001 |
| Ventricular ectopy present | 5.2 (2.4-11.3) | <0.001 |
| ST-segment abnormality | 4.7 (2.1-10.5) | <0.001 |
| Random glucose 156-164 mg/dL | 3.1 (1.4-6.8) | 0.005 |
| Heart rate >85 bpm | 1.6 (0.7-3.5) | 0.24 |

*Model adjusted for all variables listed. Reference categories: female sex, age 35-40, BMI <25, pulse pressure <50 mmHg, no ectopy, no ST changes, glucose <140 mg/dL, heart rate ≤85 bpm.*

Among female participants, the high-risk profile was less distinct, though elevated pulse pressure ≥55 mmHg and random glucose ≥145 mg/dL were associated with increased risk (combined OR 4.1, 95% CI: 1.3-13.2).

### Temporal Patterns of Risk Score Evolution

Analysis of risk score trajectories showed that true positive predictions typically emerged 3-5 weeks before clinical detection, with mean lead time of 4.2 weeks (SD 1.1 weeks). Risk scores in true positive cases showed progressive elevation over serial weekly measurements, whereas false positive cases more commonly exhibited isolated single-week elevations that did not persist.

## DISCUSSION

This prospective cohort study demonstrates that integration of readily accessible physiological biomarkers with machine learning algorithms can identify individuals at elevated near-term cardiovascular risk with high specificity (98.05%) and moderate sensitivity (71.05%) approximately 4-6 weeks before clinical detection through comprehensive cardiac evaluation. To our knowledge, this represents one of the first prospective validations of ML-based near-term (weeks) rather than long-term (years) CVD risk prediction in an asymptomatic working-age population.

### Principal Findings and Clinical Implications

The model’s high specificity and negative predictive value (97.6%) indicate that individuals receiving low-risk predictions are very unlikely to develop detectable cardiovascular pathology in the subsequent 4-6 weeks. This characteristic makes the system potentially valuable as a population-level screening tool to efficiently identify individuals who may benefit from more intensive evaluation, while providing reassurance to the majority who remain at low near-term risk. In a screening population of 500 individuals, the model correctly classified 480 (96%), substantially reducing the burden of unnecessary comprehensive cardiac workups while capturing approximately 71% of cases that ultimately developed disease.

The moderate sensitivity (71.05%), while lower than specificity, must be interpreted in the context of the study’s design and clinical use case. Missing 11 of 38 CVD cases (false negatives) represents a limitation; however, several factors mitigate this concern. First, the false negative cases tended to represent more slowly progressive or paroxysmal conditions less amenable to prediction weeks in advance. Second, in a real-world implementation, weekly screening provides multiple opportunities for detection rather than a single time-point assessment. Third, this system is intended to complement rather than replace standard clinical risk assessment and periodic comprehensive evaluation.

The positive predictive value of 75% means that three-quarters of individuals flagged as high-risk truly had or were developing cardiovascular disease. While the 25% false positive rate necessitates confirmatory evaluation, the consequences of these false alarms—additional cardiac testing that proves reassuring—are far less severe than the consequences of false negatives (missed disease). This asymmetry in clinical consequences supports adoption of a somewhat lower specificity threshold than might be acceptable for diseases where false positives carry greater harm.

### Comparison with Existing Literature

Traditional cardiovascular risk scores (Framingham, SCORE, pooled cohort equations) typically predict 10-year risk and demonstrate c-statistics of 0.70-0.80 (6, 8), comparable to or somewhat lower than our AUROC of 0.923. However, direct comparison is inappropriate given the fundamental difference in prediction horizons. Our system identifies imminent risk (4-6 weeks), enabling immediate targeted intervention, whereas traditional scores inform long-term preventive strategies.

Recent studies have explored ML approaches to cardiovascular risk prediction with variable results. Weng et al. (25) demonstrated that neural networks could outperform traditional risk scores for 10-year CVD prediction (AUROC 0.764 vs 0.728), though still limited to long-term forecasting. Khera et al. (26) showed that polygenic risk scores combined with clinical factors improved risk stratification but again focused on lifetime or 10-year horizons.

More analogous to our approach, several studies have investigated near-term risk prediction in specific contexts. Johnson et al. (27) used ML to predict 30-day risk of major adverse cardiac events in emergency department patients with chest pain, achieving similar sensitivity (68%) and higher specificity (92%) in a higher-risk population. Krittanawong et al. (28) developed deep learning models to predict cardiovascular events within 6 months from ECG data, demonstrating AUROC of 0.88 but in a population with established coronary disease rather than asymptomatic individuals.

The integration of single-lead ECG deserves particular attention. While 12-lead ECG remains the diagnostic standard, recent work has shown that single-lead devices can detect atrial fibrillation with high sensitivity (29), identify structural heart disease (30), and contribute to cardiovascular risk stratification when combined with other biomarkers (31). Our findings support the utility of single-lead ECG, particularly when ECG-derived features (ectopy, ST changes, heart rate variability) are incorporated into multimodal prediction models rather than interpreted in isolation.

### Mechanistic Insights and High-Risk Profiles

The biomarker constellation associated with highest CVD risk in our cohort—particularly the combination of elevated pulse pressure, ECG abnormalities, and elevated random glucose in males with modestly elevated BMI—aligns with established pathophysiological understanding. Pulse pressure, the difference between systolic and diastolic blood pressure, reflects arterial stiffness and is a well-established independent cardiovascular risk factor (32, 33). Widened pulse pressure indicates reduced arterial compliance, often due to subclinical atherosclerosis, and predicts cardiovascular events independent of systolic or diastolic pressure alone.

The prominence of ECG abnormalities, particularly ventricular ectopy and ST-segment changes, as risk markers warrants consideration. Frequent premature ventricular contractions, even in apparently healthy individuals, associate with increased cardiovascular and all-cause mortality (34, 35). ST-segment deviations, when persistent or progressive, may indicate subclinical ischemia or cardiomyopathy. The weekly ECG monitoring in our protocol likely captured transient abnormalities that would be missed in episodic clinic-based screening, enhancing the model’s predictive capability.

Elevated random glucose, even when not meeting diagnostic thresholds for diabetes, confers cardiovascular risk through multiple mechanisms including endothelial dysfunction, oxidative stress, and promotion of atherosclerosis (36, 37). The glucose range associated with highest risk in our study (156-164 mg/dL random) suggests pre-diabetes or early diabetes, conditions known to substantially elevate cardiovascular risk.

The sex-specific differences in risk profiles deserve attention. Male participants demonstrated higher baseline risk parameters and comprised 73.7% of CVD cases despite representing 60% of the cohort. This male predominance aligns with well-established sex differences in cardiovascular disease epidemiology, with males typically experiencing earlier onset of atherosclerotic disease (38). The less distinct high-risk profile in females may reflect different pathophysiological mechanisms, with females more commonly presenting with microvascular dysfunction and non-obstructive coronary disease (39, 40), patterns potentially less readily captured by our biomarker panel.

### Implementation Considerations and Clinical Integration

Translation of this ML system into clinical practice requires careful consideration of implementation strategies and workflow integration. Several models merit consideration:

#### Employer-Based Screening Programs

Corporate wellness initiatives could integrate weekly screening with existing occupational health services, providing convenient access for working-age adults—the population most likely to be asymptomatic despite elevated risk. Our study’s high adherence rate (98.4%) suggests feasibility when screening is conveniently located at worksites.

#### Community Health Screening

Deployment of screening kiosks in pharmacies, shopping centers, or community health centers could extend reach beyond employed populations, though maintaining engagement with weekly screening outside structured workplace programs may prove challenging.

#### Insurance-Based Prevention Programs

Health insurers seeking to reduce cardiovascular event costs could incentivize participation in monitoring programs, with risk-stratified interventions for individuals flagged as high-risk. The documented cost-effectiveness of preventing cardiovascular events (41, 42) supports the business case for such programs, particularly given this system’s low per-screening cost and high specificity.

Critical to any implementation is establishing clear protocols for responding to high-risk alerts. In our study, all risk scores were concealed during the observation period to enable unbiased outcome ascertainment. In clinical deployment, elevated risk scores should trigger structured pathways including physician notification, comprehensive clinical evaluation (ideally including 12-lead ECG, fasting lipids and glucose, and potentially advanced imaging), and initiation of appropriate preventive therapies (statins, antihypertensives, antiplatelet agents, lifestyle interventions) based on comprehensive assessment.

The 4-6 week prediction window is clinically actionable. This timeframe permits scheduling of subspecialty consultations and diagnostic testing that often require weeks to arrange, while being sufficiently near-term that identified risk likely reflects current physiological state rather than distant statistical probability. Interventions intensified in response to elevated risk scores—whether pharmacological, behavioral, or diagnostic—can realistically be implemented within this window.

### Limitations and Methodological Considerations

Several limitations warrant acknowledgment. First, the study population comprised employed adults aged 35-50 years in Pakistan, potentially limiting generalizability to other age groups, geographic regions, or populations with different baseline risk profiles. The exclusion of individuals with known cardiovascular disease, while appropriate for assessing screening utility, means performance in higher-risk populations remains unknown.

Second, the relatively small number of CVD events (n=38) limited precision of sensitivity estimates and statistical power for subgroup analyses, particularly for females who experienced only 10 events. Larger validation cohorts are needed to more precisely characterize performance across diverse subgroups and to identify whether certain populations benefit disproportionately from this screening approach.

Third, defining the outcome as any cardiovascular finding detected through comprehensive monthly evaluation differs from traditional endpoints such as myocardial infarction, stroke, or cardiovascular death. While our composite endpoint enabled prospective validation within a 12-month study period, the clinical significance of some detected abnormalities (e.g., mild LV dysfunction, asymptomatic arrhythmias) varies. Longer follow-up examining major adverse cardiac events (MACE) is needed to determine whether early detection translates into improved hard clinical outcomes.

Fourth, the monthly comprehensive evaluations, while providing gold-standard outcome ascertainment for research purposes, would be impractical and cost-prohibitive in routine clinical practice. Our validation approach may have detected CVD cases earlier than they would manifest clinically, potentially inflating apparent model performance. However, the fact that detected abnormalities required subspecialist diagnosis using advanced imaging suggests clinical relevance rather than trivial findings.

Fifth, the ML model was trained on separate retrospective data, but detailed characteristics of the training dataset, including geographic origin and clinical settings, were not extensively described herein. Differences between training and validation populations could affect generalizability. Additionally, while hyperparameters were locked prospectively, model calibration was not formally assessed beyond discrimination metrics. Future work should evaluate calibration across risk strata to ensure predicted probabilities accurately reflect observed event rates.

Sixth, the weekly screening protocol, while demonstrating feasibility in our study, may prove burdensome in routine practice, potentially affecting adherence in less structured settings. Alternative frequencies (e.g., biweekly screening) should be explored to optimize the balance between predictive accuracy and participant burden.

Finally, cost-effectiveness analysis was not performed. While avoiding cardiovascular events clearly provides value, the incremental costs of screening infrastructure, confirmatory evaluations for positive screens, and interventions must be weighed against benefits. Such analysis requires longer follow-up to capture downstream clinical events and associated costs.

### Future Directions and Model Enhancement

Several avenues for future research and model refinement merit exploration:

**1. Demographic and Geographic Generalization:** Validation in diverse populations including different age groups (younger adults, elderly), geographic regions (Western vs. Asian vs. African populations), and ethnicities is essential. Cardiovascular disease epidemiology and risk factor profiles vary substantially across populations (43, 44), and ML models may require population-specific calibration. Furthermore, incorporating genetic risk scores or ancestry-specific adjustments may reduce demographic biases and improve performance across diverse populations.
**2. Additional Biomarkers:** Integration of additional readily accessible biomarkers could enhance predictive accuracy. Candidates include point-of-care lipid measurements, natriuretic peptides (BNP/NT-proBNP), high-sensitivity troponin, and inflammatory markers (hs-CRP). Wearable device data including continuous heart rate, heart rate variability, physical activity patterns, and sleep metrics represent particularly promising additions given the increasing prevalence of consumer wearables (45, 46).
**3. Advanced ECG Analysis:** While our single-lead ECG analysis captured basic features, deep learning approaches applied directly to raw ECG waveforms have shown promise in detecting subtle patterns predictive of cardiovascular outcomes (47, 48). Integration of convolutional neural networks trained on ECG signals could potentially improve sensitivity by identifying patterns not easily quantified through conventional feature engineering.
**4. Temporal Modeling:** Our model incorporated some temporal features (rate of change in parameters), but more sophisticated approaches using recurrent neural networks (RNNs) or long short-term memory (LSTM) networks could better capture longitudinal trajectories and identify patterns of physiological deterioration over time (49, 50). Such approaches might particularly improve detection of gradually progressive conditions that were disproportionately represented among false negatives in our study.
**5. Mechanistic Phenotyping:** Rather than treating CVD as a single outcome, future work should examine whether different cardiovascular phenotypes (ischemic heart disease, heart failure, arrhythmias, valvular disease) have distinct predictive features. Disease-specific models might achieve higher performance than the composite endpoint model evaluated herein.
**6. Intervention Trials:** The ultimate validation of any screening tool is demonstration that screening-prompted interventions improve outcomes. Randomized controlled trials comparing outcomes in populations with vs. without access to ML-based risk monitoring are needed to definitively establish clinical utility and cost-effectiveness.
**7. Addressing Algorithmic Bias:** ML models can perpetuate or amplify biases present in training data, potentially leading to disparities in performance across demographic groups (51, 52). Rigorous evaluation for bias, including intersectional analyses examining performance across combinations of sex, age, and ethnicity, should be standard. Techniques for bias mitigation including balanced sampling, fairness constraints, and subgroup-specific calibration should be explored.
**8. Explainability and Clinical Adoption:** Healthcare providers are appropriately cautious about black-box algorithms that provide predictions without interpretable rationale (53). Development of explainability tools that identify which specific biomarker patterns drove individual risk predictions could facilitate clinical trust and adoption. Methods such as SHAP (SHapley Additive exPlanations) values or attention mechanisms in neural networks can provide insight into model decision-making.
**9. Multi-Disease Models:** Future iterations might predict not only cardiovascular disease but also metabolic conditions (diabetes), renal disease, and other chronic diseases sharing common risk factors, maximizing the value derived from serial biomarker monitoring.
**10. Real-World Implementation Studies:** Pragmatic trials examining implementation in real-world settings—including assessment of adherence, clinical workflow integration, provider acceptance, patient satisfaction, and health system costs—are essential for translating research findings into sustainable clinical practice.

## CONCLUSIONS

This prospective validation study demonstrates that machine learning-based integration of routine physiological parameters enables prediction of near-term (4-6 weeks) cardiovascular disease risk in asymptomatic working-age adults with high specificity (98.05%), moderate sensitivity (71.05%), and overall accuracy of 96.0%. The system’s high negative predictive value (97.6%) makes it potentially valuable for population-level screening to efficiently identify individuals warranting intensive evaluation while providing reassurance to the majority at low near-term risk.

The identification of a high-risk biomarker constellation—particularly widened pulse pressure, ECG abnormalities (ventricular ectopy, ST changes), and elevated random glucose—provides mechanistic insights and could inform targeted screening strategies even in the absence of sophisticated ML algorithms.

While promising, several limitations including modest sample size, single-region validation, and lack of hard clinical outcome data necessitate cautious interpretation. Extensive additional validation across diverse populations, longer-term follow-up to assess impact on major adverse cardiac events, and pragmatic implementation trials are needed before widespread clinical deployment can be recommended.

This work represents a step toward realizing the vision of continuous, accessible cardiovascular monitoring enabling early intervention to prevent acute events. As wearable technology and point-of-care diagnostics continue to advance, integration of serial biomarker data with sophisticated analytical methods offers the potential to fundamentally transform cardiovascular prevention from episodic risk estimation to dynamic, personalized surveillance.

## ACKNOWLEDGMENTS

We express our sincere gratitude to Dr. Abdul Hameed Bhatti, Department of Critical Care, PAF Hospital Islamabad, for his invaluable clinical expertise, guidance in study design, and oversight of cardiac outcome adjudication. His deep understanding of cardiovascular pathophysiology and dedication to preventive medicine were instrumental to this work.

We thank ProMed Solutions Pvt. Ltd. for their essential role in participant recruitment, coordination of weekly screening sessions across multiple corporate sites, and logistical support throughout the study period. Their commitment to employee health and wellness enabled the successful completion of this research.

We are deeply grateful to the leadership, cardiologists, sonographers, and laboratory staff at PAF Hospital Islamabad for conducting the comprehensive monthly cardiac evaluations that served as the gold standard for outcome ascertainment. Their meticulous attention to diagnostic quality and willingness to accommodate the intensive evaluation schedule required for this research were indispensable.

Finally, we thank the 500 study participants who generously volunteered their time over 12 months, attending weekly screenings and monthly evaluations. Their commitment to advancing cardiovascular health research made this study possible.

## DATA AVAILABILITY

De-identified individual participant data may be made available to qualified researchers upon reasonable request and subject to approval by the institutional ethics committee and execution of appropriate data use agreements. Requests should be directed to the corresponding author. The trained machine learning model will not be shared to protect proprietary intellectual property, but model architecture and hyperparameters are described in sufficient detail to enable replication.

## REFERENCES

1. Roth GA, Mensah GA, Johnson CO, et al. Global Burden of Cardiovascular Diseases and Risk Factors, 1990–2019: Update From the GBD 2019 Study. J Am Coll Cardiol. 2020;76(25):2982–3021.

2. S.S. Virani, A. Alonso, Aparicio HJ, et al. Heart Disease and Stroke Statistics—2021 Update: A Report From the American Heart Association. Circulation. 2021;143(8):e254–e743.

3. Khot UN, Khot MB, Bajzer CT, et al. Prevalence of conventional risk factors in patients with coronary heart disease. JAMA. 2003;290(7):898–904.

4. Yusuf S, Rangarajan S, Teo K, et al. Cardiovascular risk and events in 17 low-, middle-, and high-income countries. N Engl J Med. 2014;371(9):818–827.

5. D’Agostino RB Sr, Vasan RS, Pencina MJ, et al. General cardiovascular risk profile for use in primary care: the Framingham Heart Study. Circulation. 2008;117(6):743–753.

6. Goff DC Jr, Lloyd-Jones DM, Bennett G, et al. 2013 ACC/AHA guideline on the assessment of cardiovascular risk: a report of the American College of Cardiology/American Heart Association Task Force on Practice Guidelines. Circulation. 2014;129(25 Suppl 2):S49–73.

7. DeFilippis AP, Young R, Carrubba CJ, et al. An analysis of calibration and discrimination among multiple cardiovascular risk scores in a modern multiethnic cohort. Ann Intern Med. 2015;162(4):266–275.

8. Pylypchuk R, Wells S, Kerr A, et al. Cardiovascular disease risk prediction equations in 400 000 primary care patients in New Zealand: a derivation and validation study. Lancet. 2018;391(10133):1897–1907.

9. Wang TJ, Gona P, Larson MG, et al. Multiple biomarkers for the prediction of first major cardiovascular events and death. N Engl J Med. 2006;355(25):2631–2639.

10. Krittanawong C, Zhang H, Wang Z, Aydar M, Kitai T. Artificial Intelligence in Precision Cardiovascular Medicine. J Am Coll Cardiol. 2017;69(21):2657–2664.

11. Ambale-Venkatesh B, Yang X, Wu CO, et al. Cardiovascular Event Prediction by Machine Learning: The Multi-Ethnic Study of Atherosclerosis. Circ Res. 2017;121(9):1092–1101.

12. Alaa AM, Bolton T, Di Angelantonio E, Rudd JHF, van der Schaar M. Cardiovascular disease risk prediction using automated machine learning: A prospective study of 423,604 UK Biobank participants. PLoS One. 2019;14(5):e0213653.

13. Dinh A, Miertschin S, Young A, Mohanty SD. A data-driven approach to predicting diabetes and cardiovascular disease with machine learning. BMC Med Inform Decis Mak. 2019;19(1):211.

14. Zhao Y, Wood EP, Mirin N, et al. Early Detection of ST-Segment Elevated Myocardial Infarction by Artificial Intelligence With 12-Lead Electrocardiogram. Int J Cardiol. 2020;317:223–230.

15. Fuster V, Kovacic JC. Acute Coronary Syndromes: Pathology, Diagnosis, Genetics, Prevention, and Treatment. Circ Res. 2014;114(12):1847–1851.

16. Bumgarner JM, Lambert CT, Hussein AA, et al. Smartwatch Algorithm for Automated Detection of Atrial Fibrillation. J Am Coll Cardiol. 2018;71(21):2381–2388.

17. Tison GH, Sanchez JM, Ballinger B, et al. Passive Detection of Atrial Fibrillation Using a Commercially Available Smartwatch. JAMA Cardiol. 2018;3(5):409–416.

18. Hannun AY, Rajpurkar P, Haghpanahi M, et al. Cardiologist-level arrhythmia detection and classification in ambulatory electrocardiograms using a deep neural network. Nat Med. 2019;25(1):65–69.

19. Attia ZI, Noseworthy PA, Lopez-Jimenez F, et al. An artificial intelligence-enabled ECG algorithm for the identification of patients with atrial fibrillation during sinus rhythm: a retrospective analysis of outcome prediction. Lancet. 2019;394(10201):861–867.

20. Yusuf S, Hawken S, Ounpuu S, et al. Effect of potentially modifiable risk factors associated with myocardial infarction in 52 countries (the INTERHEART study): case-control study. Lancet. 2004;364(9438):937–952.

21. Whelton PK, Carey RM, Aronow WS, et al. 2017 ACC/AHA/AAPA/ABC/ACPM/AGS/APhA/ASH/ASPC/NMA/PCNA Guideline for the Prevention, Detection, Evaluation, and Management of High Blood Pressure in Adults. J Am Coll Cardiol. 2018;71(19):e127–e248.

22. Coutinho M, Gerstein HC, Wang Y, Yusuf S. The relationship between glucose and incident cardiovascular events. A metaregression analysis of published data from 20 studies of 95,783 individuals followed for 12.4 years. Diabetes Care. 1999;22(2):233–240.

23. Levitan EB, Song Y, Ford ES, Liu S. Is nondiabetic hyperglycemia a risk factor for cardiovascular disease? A meta-analysis of prospective studies. Arch Intern Med. 2004;164(19):2147–2155.

24. Surawicz B, Childers R, Deal BJ, et al. AHA/ACCF/HRS recommendations for the standardization and interpretation of the electrocardiogram. J Am Coll Cardiol. 2009;53(11):976–981.

25. Weng SF, Reps J, Kai J, Garibaldi JM, Qureshi N. Can machine-learning improve cardiovascular risk prediction using routine clinical data? PLoS One. 2017;12(4):e0174944.

26. Khera AV, Chaffin M, Aragam KG, et al. Genome-wide polygenic scores for common diseases identify individuals with risk equivalent to monogenic mutations. Nat Genet. 2018;50(9):1219–1224.

27. Johnson KW, Torres Soto J, Glicksberg BS, et al. Artificial Intelligence in Cardiology. J Am Coll Cardiol. 2018;71(23):2668–2679.

28. Krittanawong C, Virk HUH, Bangalore S, et al. Machine learning prediction in cardiovascular diseases: a meta-analysis. Sci Rep. 2020;10(1):16057.

29. Perez MV, Mahaffey KW, Hedlin H, et al. Large-Scale Assessment of a Smartwatch to Identify Atrial Fibrillation. N Engl J Med. 2019;381(20):1909–1917.

30. Attia ZI, Kapa S, Lopez-Jimenez F, et al. Screening for cardiac contractile dysfunction using an artificial intelligence-enabled electrocardiogram. Nat Med. 2019;25(1):70–74.

31. Raghunath S, Pfeifer JM, Ulloa-Cerna AE, et al. Deep Neural Networks Can Predict New-Onset Atrial Fibrillation From the 12-Lead ECG and Help Identify Those at Risk of Atrial Fibrillation-Related Stroke. Circulation. 2021;143(13):1287–1298.

32. Mitchell GF, Hwang SJ, Vasan RS, et al. Arterial stiffness and cardiovascular events: the Framingham Heart Study. Circulation. 2010;121(4):505–511.

33. Safar ME, Levy BI, Struijker-Boudier H. Current perspectives on arterial stiffness and pulse pressure in hypertension and cardiovascular diseases. Circulation. 2003;107(22):2864–2869.

34. Dukes JW, Dewland TA, Vittinghoff E, et al. Ventricular Ectopy as a Predictor of Heart Failure and Death. J Am Coll Cardiol. 2015;66(2):101–109.

35. Ataklte F, Erqou S, Laukkanen J, Kaptoge S. Meta-analysis of ventricular premature complexes and their relation to cardiac mortality in general populations. Am J Cardiol. 2013;112(8):1263–1270.

36. Emerging Risk Factors Collaboration. Diabetes mellitus, fasting blood glucose concentration, and risk of vascular disease: a collaborative meta-analysis of 102 prospective studies. Lancet. 2010;375(9733):2215–2222.

37. Sarwar N, Gao P, Seshasai SR, et al. Diabetes mellitus, fasting blood glucose concentration, and risk of vascular disease: a collaborative meta-analysis of 102 prospective studies. Lancet. 2010;375(9733):2215–2222.

38. Maas AH, Appelman YE. Gender differences in coronary heart disease. Neth Heart J. 2010;18(12):598–602.

39. Pepine CJ, Ferdinand KC, Shaw LJ, et al. Emergence of Nonobstructive Coronary Artery Disease: A Woman’s Problem and Need for Change in Definition on Angiography. J Am Coll Cardiol. 2015;66(17):1918–1933.

40. Merz CNB, Pepine CJ, Walsh MN, Fleg JL. Ischemia and No Obstructive Coronary Artery Disease (INOCA): Developing Evidence-Based Therapies and Research Agenda for the Next Decade. Circulation. 2017;135(11):1075–1092.

41. Pandya A, Weinstein MC, Gaziano TA. A comparative assessment of non-laboratory-based versus commonly used laboratory-based cardiovascular disease risk scores in the NHANES III population. PLoS One. 2011;6(5):e20416.

42. Karmali KN, Lloyd-Jones DM, van der Leeuw J, et al. Blood pressure-lowering treatment strategies based on cardiovascular risk versus blood pressure: A meta-analysis of individual participant data. PLoS Med. 2018;15(3):e1002538.

43. Yusuf S, Rangarajan S, Teo K, et al. Cardiovascular risk and events in 17 low-, middle-, and high-income countries. N Engl J Med. 2014;371(9):818–827.

44. Joshi P, Islam S, Pais P, et al. Risk factors for early myocardial infarction in South Asians compared with individuals in other countries. JAMA. 2007;297(3):286–294.

45. Quer G, Radin JM, Gadaleta M, et al. Wearable sensor data and self-reported symptoms for COVID-19 detection. Nat Med. 2021;27(1):73–77.

46. Turakhia MP, Desai M, Hedlin H, et al. Rationale and design of a large-scale, app-based study to identify cardiac arrhythmias using a smartwatch: The Apple Heart Study. Am Heart J. 2019;207:66–75.

47. Ribeiro AH, Ribeiro MH, Paixão GMM, et al. Automatic diagnosis of the 12-lead ECG using a deep neural network. Nat Commun. 2020;11(1):1760.

48. Attia ZI, Noseworthy PA, Lopez-Jimenez F, et al. An artificial intelligence-enabled ECG algorithm for the identification of patients with atrial fibrillation during sinus rhythm: a retrospective analysis of outcome prediction. Lancet. 2019;394(10201):861–867.

49. Choi E, Schuetz A, Stewart WF, Sun J. Using recurrent neural network models for early detection of heart failure onset. J Am Med Inform Assoc. 2017;24(2):361–370.

50. Rajkomar A, Oren E, Chen K, et al. Scalable and accurate deep learning with electronic health records. NPJ Digit Med. 2018;1:18.

51. Obermeyer Z, Powers B, Vogeli C, Mullainathan S. Dissecting racial bias in an algorithm used to manage the health of populations. Science. 2019;366(6464):447–453.

52. Gianfrancesco MA, Tamang S, Yazdany J, Schmajuk G. Potential Biases in Machine Learning Algorithms Using Electronic Health Record Data. JAMA Intern Med. 2018;178(11):1544–1547.

53. Holzinger A, Biemann C, Pattichis CS, Kell DB. What do we need to build explainable AI systems for the medical domain? arXiv:1712.09923 [cs.AI]. 2017.

